# Combined visualization of genomic and epidemiological data for outbreaks

**DOI:** 10.1101/2024.04.02.24305229

**Authors:** Carl J.E. Suster, Anne E. Watt, Qinning Wang, Sharon C.-A. Chen, Jen Kok, Vitali Sintchenko

## Abstract

In epidemiological investigations, pathogen genomics can provide insights and test epidemiological hypotheses that would not have been possible through traditional epidemiology. Tools to synthesize genomic analysis with other types of data are a key requirement of genomic epidemiology. We propose a new ‘phylepic’ visualization that combines a phylogenomic tree with an epidemic curve. The combination visually links the molecular time represented in the tree to the calendar time in the epidemic curve, a correspondence that is not easily represented by existing tools. Using an example derived from a foodborne bacterial outbreak, we demonstrated that the phylepic chart communicates that what appeared to be a point-source outbreak was in fact composed of cases associated with two genetically distinct clades of bacteria. We provide an R package implementing the chart. We expect that visualizations that place genomic analyses within the epidemiological context will become increasingly important for outbreak investigations and public health surveillance of infectious diseases.

---

For investigations of infectious disease outbreaks, high-resolution characterisation and clustering of pathogen genomes using whole genome sequencing (WGS) has proven to be a powerful complementary tool to epidemiological investigations [1,2]. A epidemiological investigation will often involve establishing a case definition to demarcate the outbreak, geospatial and temporal clustering, case follow-up and further investigations to establish transmission pathways or common reservoirs [3]. Genomic epidemiology draws together bioinformatic analysis from pathogen genomic sequences and epidemiological data to provide context and support to inferences of transmission. Synthesis of genomic and epidemiological data is required to perform these inferences, however this can be challenging given the complexity of the underlying data [4]. These challenges can be compounded when genomic and epidemiological investigations are conducted by discrete organisations (for example public health laboratories and local public health services respectively) with different information contexts and objectives.

The epidemic curve, a histogram of cases binned along the time axis, is a key representation relied upon in disease control. It is straightforward to prepare from line listing data, supports both data exploration and statistical epidemiology [5], and is straightforward to interpret. Genomic epidemiology instead relies upon phylogenomic trees, which display a detailed summary of the relationships between the included genomes under the particular assumptions of the underlying evolutionary models. Phylogenomic trees are prepared using specialized tools that implement the evolutionary models and efficiently search for the optimal tree (e.g. IQ-TREE, http://www.iqtree.org/). Trees are an essential representation of the salient information contained in a set of genomes that support genomic epidemiology and public health surveillance. Visually extracting this information relies upon knowledge of the conventions of tree drawing, and some familiarity with the process of their preparation. Users of trees may therefore require dedicated training so as to minimize the risk of misinterpretation.

We propose a new visualization called the ‘phylepic’ chart that synthesizes epidemic curves and phylogenomic trees. The structure of the phylepic chart facilitates visual linkage of the molecular time represented in the tree with the epidemiological time represented in the epidemic curve [6]. For illustration, we adapted a dataset from the investigation of a foodborne point-source outbreak in New South Wales (NSW), Australia. Bacterial isolates were collected during the outbreak period and underwent WGS. The resulting sequences were analysed alongside historical sequences collected in NSW using conventional bioinformatic pipelines to produce a core genome phylogenomic tree. For the purpose of illustration, we have pruned the resulting phylogeny and added random noise to the dates associated with each isolate. The chart is presented in Figure 1.

**Figure 1.**
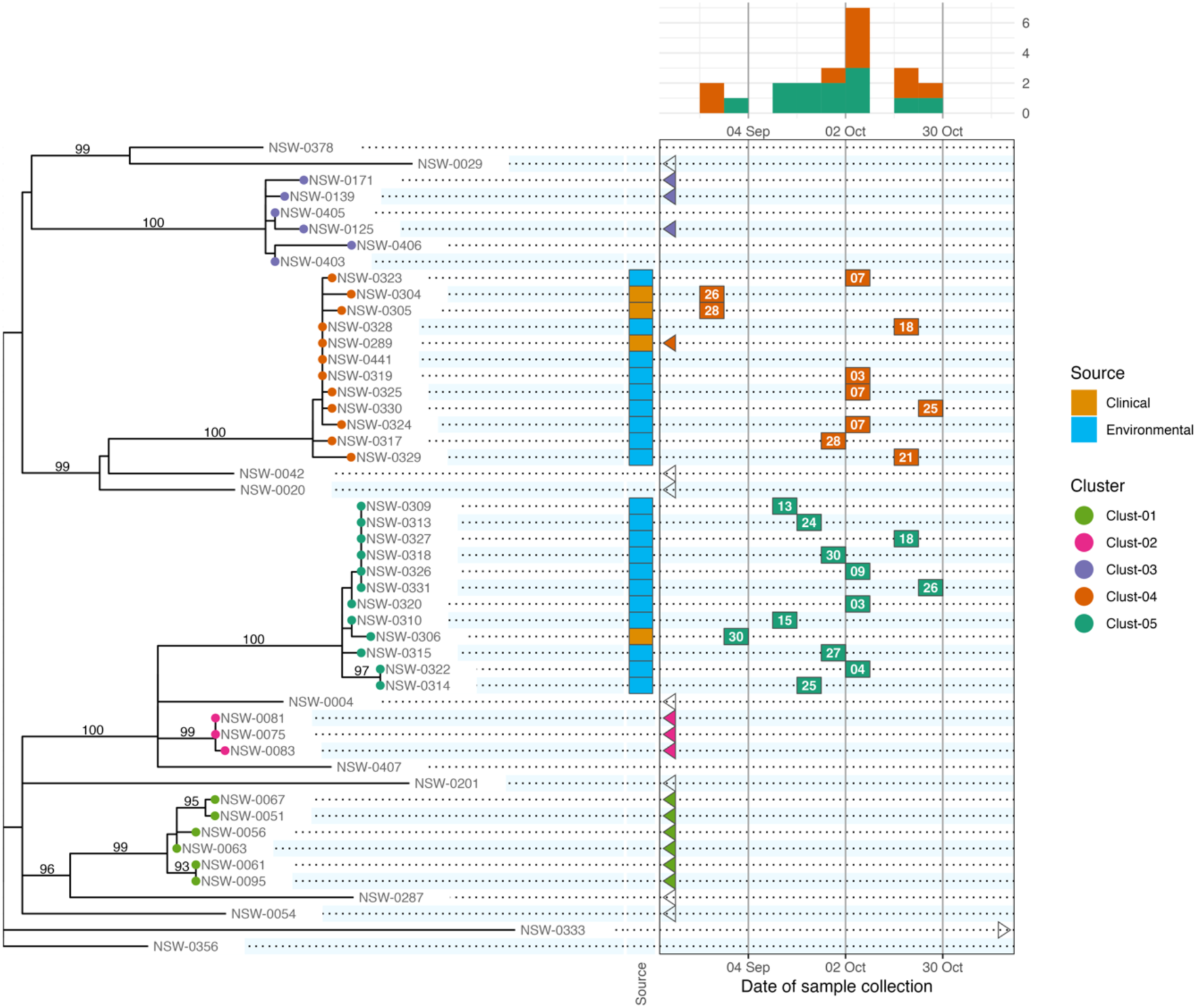
A phylepic chart using an illustrative dataset adapted from a foodborne enteric pathogen outbreak in NSW. The panel on the left shows the phylogenomic tree with tips colours by genomic cluster membership. Numbers on branches are bootstrap support values expressed as a percentage. The panel on the upper right shows the epidemic curve using date of sample collection as the time for each isolate. The calendar grid (boxed) in the lower right connects the tree to the epidemic curve, and on each tile shows the day of the month of the sample collection date. Samples with a collection date outside the outbreak period are shown with triangles at the closest edge of the date scale. Colours on tree tips, calendar tiles, and the epidemic curve refer to the same genomic clusters.

Relying on the epidemiological data alone, the silhouette of the epidemic curve in Figure 1 is consistent with a single point-source outbreak. The epidemic curve is coloured by genomic cluster membership. The presence of two genomic clusters suggests that there are two distinct strains associated with cases occurring in the same period. The phylogenomic tree provides more complete genomic context of the outbreak by providing detailed information about the inferred relatedness of bacterial sequences. The tree reveals that the most recent common ancestor of the two outbreak clusters (Clust-04 and Clust-05) is also the common ancestor of several other clusters and singletons in the historical dataset. This provides evidence that Clust-04 and Clust-05 arose from separate introductions of the pathogen. From the tree alone the outbreak is not evident as there is no representation of the temporal proximity of cases. Only the combination of the two lines of evidence leads to the conclusion of distinct introductions.

The calendar grid in the lower right of Figure 1 shows a visual projection of the tree onto a time axis representing the date. For this particular chart the time axis was restricted to the outbreak period, meaning that for isolates collected before mid-August there is no calendar tile. To distinguish these from cases with missing metadata, triangles are drawn at the edges of the calendar to indicate out-of-range values. The epidemic curve and calendar grid are binned by epidemiological week. The phylepic chart is most readable when the tree is relatively small and there are not too many date bins, both of which are reasonable restrictions for the intended application in outbreak investigation. The bin width can be decreased to a single day or increased to another interval depending on the appropriate resolution for an investigation.

While we have chosen a foodborne bacterial pathogen, phylepic charts are equally compatible with any other pathogens such as respiratory viruses. The tree need not necessarily be built from the consensus genome or a core genome as in our example; it is sufficient that a tree of some sort is prepared that is meaningful in the context of the investigation. It remains important to convey any caveats and limitations of this analysis alongside the results to ensure correct interpretation.

To communicate relevant epidemiological context, most popular tools for visualizing trees (e.g. the ggtree R package, https://yulab-smu.top/treedata-book/, or the ETE toolkit Python library, http://etetoolkit.org/) allow tips to be annotated with an array of coloured boxes or symbols. This can work well for categorical data or binary such as the isolation source of samples, results of laboratory assays, or presence of antimicrobial resistance, toxins, or virulence factors. These annotations can be incorporated into the phylepic chart, as shown with the coloured tiles indicating clinical and environmental sample sources in Figure 1. Additional columns can be incorporated to represent other categorical or quantitative data as required. With some care, it is possible to use existing software libraries such as those mentioned above to produce a chart similar to Figure 1.

Dates associated with each case (reporting, sample collection, symptom onset) are key data for generating epidemiological hypotheses. These dates do not directly relate to the branch lengths in phylogenomic trees. Instead, the axis extending from the root of the tree to its tips can be thought of as molecular time, reflecting the evolutionary distances predicted by the model. It is possible to express branch lengths in units of time using an inferred molecular clock rate, and by incorporating temporal information as explicit constraints in the model [7]. With a time-scaled tree of this sort, an alternative visualization is made possible where an epidemic curve is vertically aligned with the tree [6]. This can be helpful to show how population-level dynamics over a longer period correspond to structural features of the tree, however it is less suitable for high resolution outbreak investigation, since the dates associated with individual cases are not shown.

We have implemented the phylepic chart in an R package. Our package uses the ggplot2 library [8] as a framework, the ggraph library (https://ggraph.data-imaginist.com) for drawing the tree, and the cowplot library (https://wilkelab.org/cowplot) for aligning panels. We have designed the package so that minimal code is required to implement simple visualizations such as Figure 1, while full customisation and annotation of the plots is possible using the underlying frameworks. It takes as input a prebuilt phylogeny (in any commonly used tree format such as Newick) and tabular data containing at minimum a column of dates associated with each sequence in the tree. Compared to existing software libraries for annotating phylogenetic trees, the package implements routines for date handling (including automatic binning by week) and ensuring data matching and consistency across panels.

As genomic epidemiology becomes a routine part of public health outbreak investigations, effective integration of genomic and epidemiological evidence to support public health and clinical responses remains a key challenge. Communication and visualization tools will assist in conveying complex genomic data into investigation and response contexts [9,10]. Our experience with using visualizations similar to phylepic charts in public health reporting has shown that they can help epidemiologists and other public health professionals to navigate phylogenomic trees by relating genomic features to more familiar details of the cases under investigation.

## Data Availability

All data and software used to generate the figure in the manuscript are available at https://zenodo.org/doi/10.5281/zenodo.10906141

## Acknowledgements

We gratefully acknowledge Health Protection NSW and the NSW Health Pathology-ICPMR Microbial Genomics Reference Laboratory for providing the tree used in our example and the motivating context from the associated outbreak investigation.

## Data Availability Statement

The R package described in this report is available for download from GitHub (https://github.com/cidm-ph/phylepic) and is archived on Zenodo (https://zenodo.org/doi/10.5281/zenodo.10906141) The package also contains the tree in Newick format, metadata table in CSV format, and the R code necessary to reproduce Figure 1.

## Declarations

### Contributions

C.S.: Data Curation, Software, Visualization, Writing – original draft; A.E.W., Q.W., S.C.-A.C., J.K.: Writing – review & editing; V.S.: Conceptualization, Supervision, Writing – review & editing.

